# Phase 2 randomised placebo-controlled trial of spironolactone and dexamethasone versus dexamethasone in COVID-19 hospitalised patients in Delhi

**DOI:** 10.1101/2022.07.01.22277163

**Authors:** Bharti Wadhwa, Vikas Malhotra, Sukhyanti Kerai, Farah Husain, Nalini Bala Pandey, Kirti N Saxena, Vinay Singh, Tom M Quinn, Feng Li, Erin Gaughan, Manu Shankar-Hari, Bethany Mills, Jean Antonelli, Annya Bruce, Keith Finlayson, Anne Moore, Kevin Dhaliwal, Christopher Edwards

## Abstract

**Background:** In this phase 2 randomised placebo-controlled clinical trial, we hypothesised that blocking mineralocorticoid receptors with spironolactone in patients with COVID-19 is safe and may reduce illness severity.

**Methods:** Hospitalised patients with confirmed COVID-19 were randomly allocated to low dose oral spironolactone (50mg day 1, then 25mg once daily for 21 days) or standard care in a 2:1 ratio. Both groups received dexamethasone 6mg for 10 days. Group allocation was blinded to the patient and research team. Primary outcomes were time to recovery, defined as the number of days until patients achieved WHO Ordinal Scale (OS) category ≤ 3, and the effect of spironolactone on aldosterone, D-dimer, angiotensin II and Von Willebrand Factor (VWF).

**Results:** 120 patients were recruited in Delhi from 01 February to 30 April 2021. 74 were randomly assigned to spironolactone and dexamethasone (SpiroDex), and 46 to dexamethasone alone (Dex). There was no significant difference in the time to recovery between SpiroDex and Dex groups (SpiroDex median 4.5 days, Dex median 5.5 days, p = 0.055). SpiroDex patients had lower aldosterone levels on day 7 and lower D-dimer levels on days 4 and 7 (day 7 D-dimer mean SpiroDex 1.15µg/mL, Dex 3.15 µg/mL, p = 0.0004). There was no increase in adverse events in patients receiving SpiroDex. *Post hoc* analysis demonstrated reduced clinical deterioration (pre specified as escalating to WHO OS category >4) in the SpiroDex group vs Dex group (5.4% vs 19.6%).

**Conclusion:** Low dose oral spironolactone in addition to dexamethasone was safe and reduced D-Dimer and aldosterone. Although time to recovery was not significantly reduced, fewer patients progressed to severe disease. Phase 3 randomised controlled trials with spironolactone should be considered.

## Introduction

Most individuals infected with severe acute respiratory syndrome coronavirus-1 (SARS-CoV-2), experience mild symptoms, however the progression to severe COVID-19 is characterised by multi-organ failure, and a high risk of death (1, 2). Identifying treatments to prevent clinical deterioration is imperative.

SARS-CoV-2 requires the Angiotensin Converting Enzyme 2 (ACE2) receptor, and transmembrane serine protease 2 (TMPRSS2) on the cell membrane to facilitate cell entry (3, 4). Transcription of the TMPRSS2 gene is enhanced by androgens through androgen receptors (AR) (5), and anti-androgen treatments such as enzalutamide have been shown to reduce cellular entry of SARS-CoV-2 into human lung cells in *in vitro* studies (6). Down regulation of the ACE2 receptor as a consequence of viral infection may result in the loss of angiotensin II conversion to angiotensin (7). The resultant high levels of unopposed angiotensin II, stimulate NADPH oxidase with consequent elevation of Reactive Oxygen Species (ROS). This results in the loss of the specificity of the Mineralocorticoid Receptor (MR), enabling stimulation by cortisol and aldosterone (8). We hypothesised that spironolactone (which is a MR antagonist with anti-androgen and anti-inflammatory effects) in combination with dexamethasone, which suppresses cortisol secretion, could reduce time to recovery and severity of COVID-19 disease in hospitalised patients (6, 8-10). This hypothesis is supported by results from retrospective and prospective non-randomised open-label studies of mineralocorticoid (MR) blockade in COVID-19 patients (8, 11, 12).

In this context, we report a phase 2 randomised placebo-controlled trial (RCT) examining the addition of low dose spironolactone (SpiroDex) to standard of care (SoC), which included treatment with Dexamethasone (Dex) in a hospital setting in Delhi, India.

## Methods

### Trial Design and Patients

The trial was a single centre RCT. Patients were recruited between 01 February to 30 April 2021 from Maulana Azad Medical College and Lok Nayak Hospital, New Delhi. Patients were recruited if they had positive nasopharyngeal SARS-CoV-2 polymerase chain reaction (PCR) and/or had chest radiographic changes consistent with COVID-19 pneumonia, with an additional oxygen requirement (oxygen saturations (SpO2) <94 % on room air but maintaining SpO2 of ≥ 94 % on supplemental oxygen by mask or nasal prongs). Exclusion criteria were: participation in another clinical trial of an investigational medicinal product (IMP); mechanical ventilation; known hypersensitivity to the IMP; significant electrolyte disturbance (Hyperkalaemia: K^+^ >5.5 mmol/L); receiving potassium sparing diuretics that could not be reasonably withheld; acute renal insufficiency; if in the investigator’s opinion the patient was unwilling or unable to comply with drug administration plan; required laboratory tests or other trial procedures; pregnancy or lactating, or if the patient was on the following drugs: ACE inhibitors, amiloride, hydrocortisone, prednisolone, methylprednisolone, triamterene.

A sample size calculation was based on a primary biomarker endpoint (D-dimer) and powered to show a 60% change in treatment group compared to controls. Accepting a type 1 error rate of 0.05 the study required 108 patients at a 2 to 1 allocation providing 80% power with a 20% drop out rate. For study feasibility reasons a total of 120 patients was chosen.

### Randomisation and blinding

2:1 (SpiroDex:Dex) randomisation was performed utilising a coin toss method to achieve the desired probability The group allocation was known only to the treating non-trial physician. The patients and the trial investigators were blinded to group allocation.

### Interventions

Patients randomised to the SpiroDex group received oral spironolactone 50 mg once daily (as two tablets of 25mg) on day 1, followed by 25 mg once daily until day 21. All patients received SoC which included supportive measures for SARS-CoV-2 and approved therapies at the time as per Government of India guidelines. Dexamethasone was given from day one at a dose of 3 mg twice daily for ten days. The control arm received two placebo tablets on day one, followed by one placebo tablet daily until day 21.

In the event of hospital discharge, dexamethasone was continued until day 10 in both groups. Spironolactone 25 mg was continued for a total period of 21 days in the trial group. Placebo tablets continued until day 21 in the control arm. All patients were followed up daily by telephone to day 28 from the day of enrolment.

If a National Early Warning Score (NEWS2) score (13) became > 7 or WHO ordinal scale (WHO OS) (14) > 4 (hospitalised severe disease), the patient was withdrawn due to clinical deterioration and documented as having met the endpoint of clinical deterioration. No further clinical data was recorded thereafter.

### Clinical and Laboratory Monitoring

Baseline investigations were performed including full blood count, blood sugar, liver function tests, blood urea, serum creatinine, serum electrolytes, D-dimer, electrocardiogram (ECG), and chest X-ray (CXR). All patients were observed and vital signs monitored daily (blood pressure, heart rate, temperature, oxygen saturation (SpO2), respiratory rate, NEWS2 score. The change in oxygen saturation to fraction of inspired oxygen ratio (SpO2/FiO2) was measured daily from the start of the trial until day 10 or hospital discharge. The duration of total hospital stay as well as the duration of oxygen use, and the time in days taken to achieve an SpO2 of ≥94% on room air were recorded in both groups. Self-reported cough symptom scores were recorded for all in-patients using an ordinal scale until day 10 or until patient discharge, and again at day 28 by follow-up phone call.

### Outcomes

The primary outcomes were time to recovery, (as defined as the first day after enrolment, where the patient met the criteria of WHO OS score ≤ 3); and circulating Von Willebrand Factor (VWF), angiotensin II, aldosterone, and D-dimer levels taken at baseline, day 4 and day 7.

The secondary outcomes were to study the effect of spironolactone on the patient’s cough, determine the duration (in days) of oxygen use and oxygen-free days, and to determine the effect of spironolactone on plasma corticosteroid levels as a measure of suppression of the production of cortisol and blockade of the MR.

### Blood biomarker measurement

Patient serum was drawn on day 1, 4 and 7 to measure angiotensin II, cortisone, cortisol and aldosterone. Patient citrate plasma was used to measure VWF and D-Dimer. All assays were performed according to the manufacturer’s instructions. Concentrations were calculated against standard curves and used for statistical analysis. Blood glucose was measured by glucometer (Glucocare Sense, RMD Mediaids ltd, Haryana, India) and sodium, urea and potassium were measured as part of routine clinical care.

The circulating blood biomarkers were assayed using the following kits: Angiotensin II Enzyme-linked Immunosorbent Assay (ELISA) (E13652287, Sincere Biotech Co.,Ltd, New Taipei City 221, Taiwan (R.O.C.)); Cortisone ELISA (E13652408, Sincere Biotech Co.,Ltd, Sincere Biotech Co.,Ltd, New Taipei City 221, Taiwan (R.O.C.)); Architect Cortisol chemiluminescent microparticle immunoassay (CMIA) (8D15-25, Abbott Laboratories, Abbott Park, Illinois, USA); Liaison Aldosterone chemiluminescent immunoassay (CLIA) (310450, DiaSorin, Sallugia, Italy); Innovance VWF:Ac (10487040/OPHL03, Siemens Healthcare Diagnostics, Marburg, Germany); and Tina-quant D-Dimer Gen.2 particle-enhanced immunoturbidimetric assay (04912551, Roche Diagnostics GmbH, Mannheim, Germany).

### Statistics

A blinded database was created in MS-EXCEL by the Delhi team. Two UK investigators assessed the data independently. Data was analysed with GraphPad Prism version 9. Following completion of data processing, the trial was un-blinded. To assess time to WHO OS ≤ 3, daily WHO OS were analysed by simple linear regression and two-way ANOVA (withdrawn patients who failed to reach WHO OS ≤ 3 were excluded from the analysis). Blood biomarkers were analysed via two-way ANOVA. Chi-square tests were applied to assess the difference between proportions for categorical variables, and the two-way ANOVA, Student’s t-test, and/or simple linear regression were used for assessing significance in the difference of means for continuous variables, as appropriate. The Mann-Whitney test was used where measurements followed non-Gaussian spread. A p-value <0.05 was considered statistically significant.

### Ethical statement

Ethical approval was granted by the Institutional Ethics Committee Maulana Azad Medical College & associated Lok Nayak on 04/01/2021-code F.1/IEC/MAMC/82/10/2020/No.300.

### Registration

The trial was registered on the Clinical Trials Registry of India TRI: CTRI/2021/03/031721, reference: REF/2021/03/041472.

## Results

### Demographics

A total of 158 hospital inpatients were screened for inclusion (Figure 1). 38 were excluded due to not meeting inclusion criteria or patient choice. 46 patients were randomised to receive SoC (Dex), and 74 patients were randomised to receive SpiroDex. The demographics for all patients are summarised in Table 1.

**Table 1.** Overview of patients enrolled to study and demographics at time of enrolment.

|  | **Total**<br>**(n= 120)** | **Standard of Care**<br>**(Dex)**<br>**(n=46)** | **Standard of Care**<br>**plus Spironolactone**<br>**(SpiroDex)**<br>**(n=74)** |
| --- | --- | --- | --- |
| **Age (mean, [SD])** | 48.8 [14.3] | 49.1 [13.9] | 48.6 [14.6] |
| **Gender** | Male: 61.7% (n= 74)<br>Female: 38.3% (n= 46) | Male 67.4% (n= 31)<br>Female 32.6% (n= 15) | Male 58.1% (n= 43)<br>Female 41.9% (n= 31) |
| **Past Medical History** |  |  |  |
| **Diabetes Mellitus** | 34.2% (n= 41) | 37.0% (n= 17) | 32.4% (n= 24) |
| **Hypertension** | 28.3% (n= 34) | 21.7% (n= 10) | 32.4% (n= 24) |
| **Coronary artery disease** | 5.8% (n= 7) | 8.7% (n= 4) | 4.1% (n= 3) |
| **COPD** | 1.7% (n= 2) | (n= 0) | 2.7% (n= 2) |
| **Asthma** | 3.3% (n=4) | (n= 0) | 5.4% (n= 4) |
| **Chronic Kidney Disease** | 0.8% (n=1) | (n= 0) | 1.4% (n= 1) |
| **CVA** | 0.8% (n= 1) | (n= 0) | 1.4% (n= 1) |
| **Cancer** | 0.8% (n= 1) | 2.2% (n= 1) | (n= 0) |
| **Baseline Clinical Findings** |  |  |  |
| **NEWS2 score Day 1<sup>#</sup>**<br>**(median)** | 3 | 4 | 3 |
| **WHO OS score Day 1<sup>#</sup>**<br>**(median)** | 4 | 4 | 4 |
| **ISARIC-4C score Day 1<sup>#</sup>**<br>**(mean [SD])** | 8.0 [3.1] | 8.391 [3.09] | 7.743 [3.11] |
| **SpO2/FiO2 ratio Day 1<sup>#</sup>**<br>**(mean [SD])** | 181.9 [61.1] | 163.0 [58.3] | 193.7 [60.3] |
| **D-Dimer µg/mL day 1<sup>#</sup>**<br>**(mean [SD])** | 1.35 [1.97] | 1.798 [2.41] | 1.062 [1.58] |

**Figure 1.**
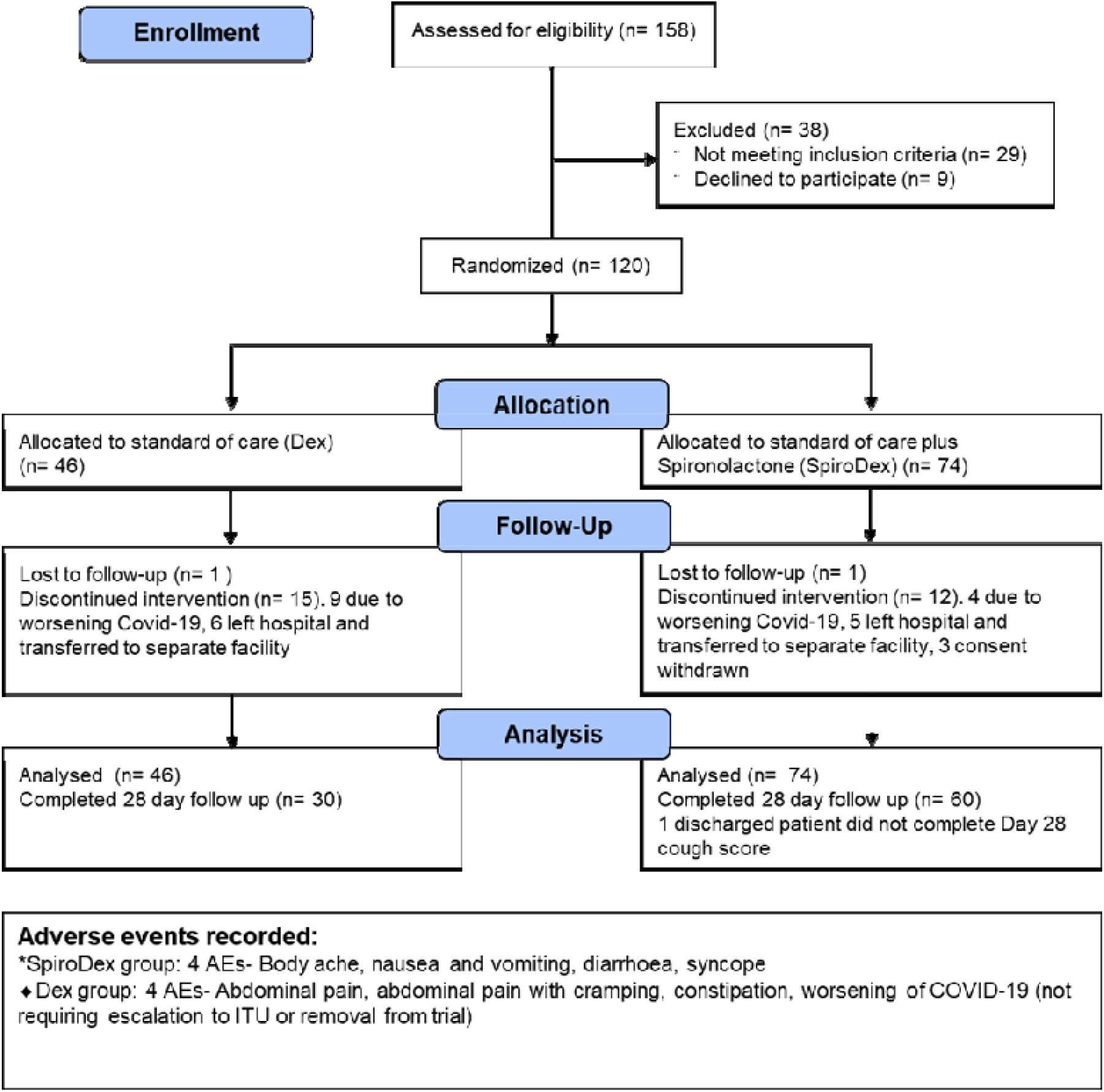
Consort diagram and adverse events for both groups.

6.8% (n = 5) of the SpiroDex arm were withdrawn as they voluntarily transferred to a separate hospital facility, compared to 13.0% (n=6) of patients in the Dex arm. These patients have been included in the analysis up until transfer from the Lok Nayak facility. The clinical outcomes for these patients after withdrawal is unknown.

### Primary outcomes

There were no statistical differences between the Dex and SpiroDex groups for the time in days for the patient to achieve WHO OS category 1, 2 or 3 (Figure 2a). The Dex group had a median of 5.5 days, compared to a median of 4.5 days for the SpiroDex group (interquartile range (IQR) Dex 2.25 days, SpiroDex 3 days, p = 0.055). In addition, there was no significant difference in the rate of change for daily WHO OS or NEWS2 measurements for these patients (Figure 2b,c). The blood biomarker results demonstrated that there was significantly reduced plasma D-dimer in the SpiroDex treatment group at days 4 and 7 compared to the Dex group (Figure 3a) (95% CI 0.77 to 3.2, p = 0.0004). VWF release was similar between both groups at days 1, 4 and 7 (Figure 3b). Aldosterone levels were significantly lower in day 7 in the SpiroDex group compared to the Dex arm (95% CI 1.65 to 13.85, p = 0.0075) (Figure 3c). Angiotensin II remained comparable across both groups (Figure 3d).

**Figure 2.**
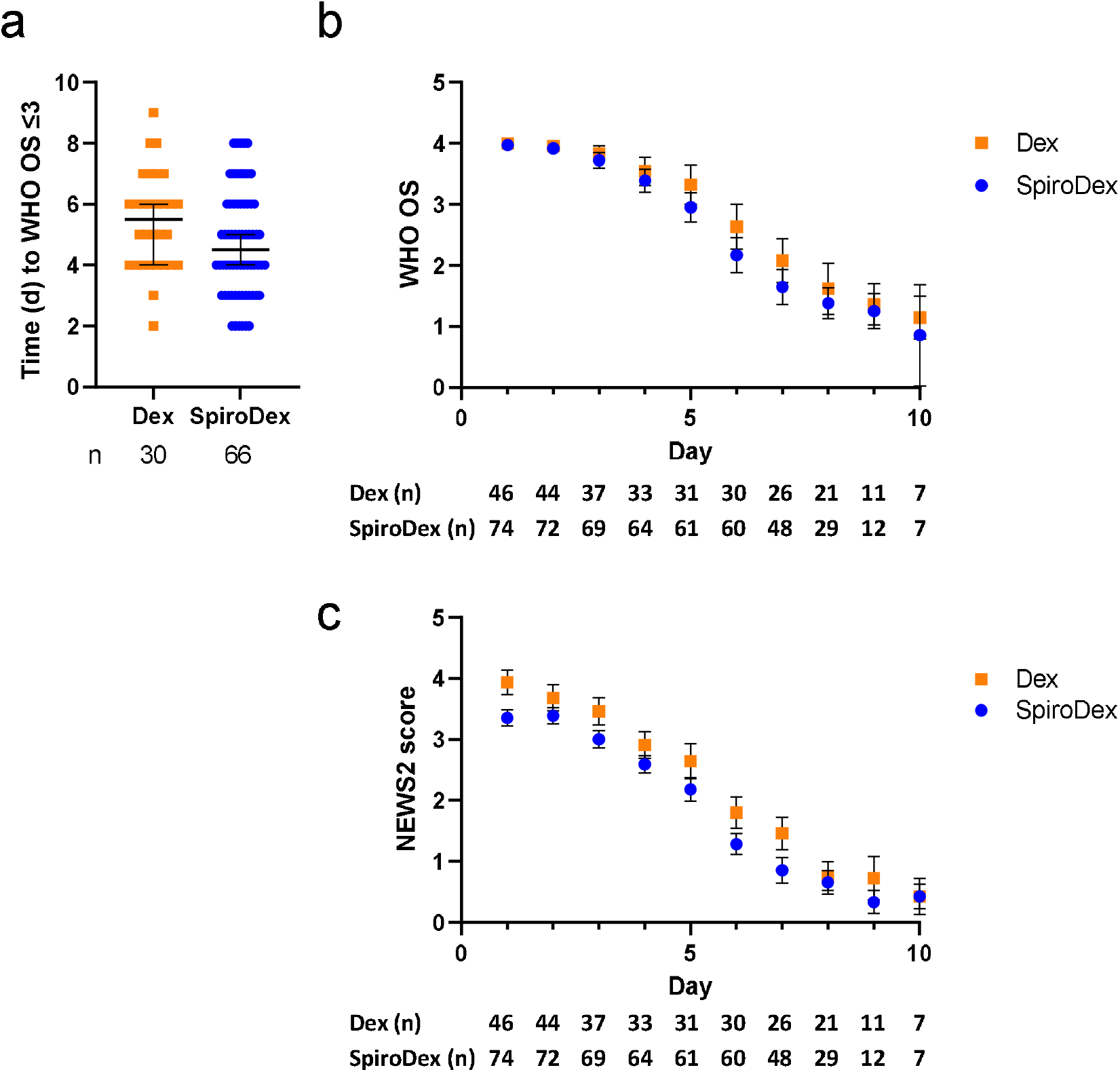
Primary Clinical Outcomes. **(a)** Time in days for patients to meet WHO OS ≤3. Median and 95% CI shown. Analysed by Mann-Whitney, p >0.05. **(b)** Daily WHO OS measurement for all patients who met WHO ordinal scale ≤3. Mean and 95% CI shown. **(c)** Daily NEWS2 score for all patients. Mean and 95% CI shown. Patient number (n) included in analysis is shown below the plot. For all, Dex: squares, orange; SpiroDex: circles,blue.

**Figure 3:**
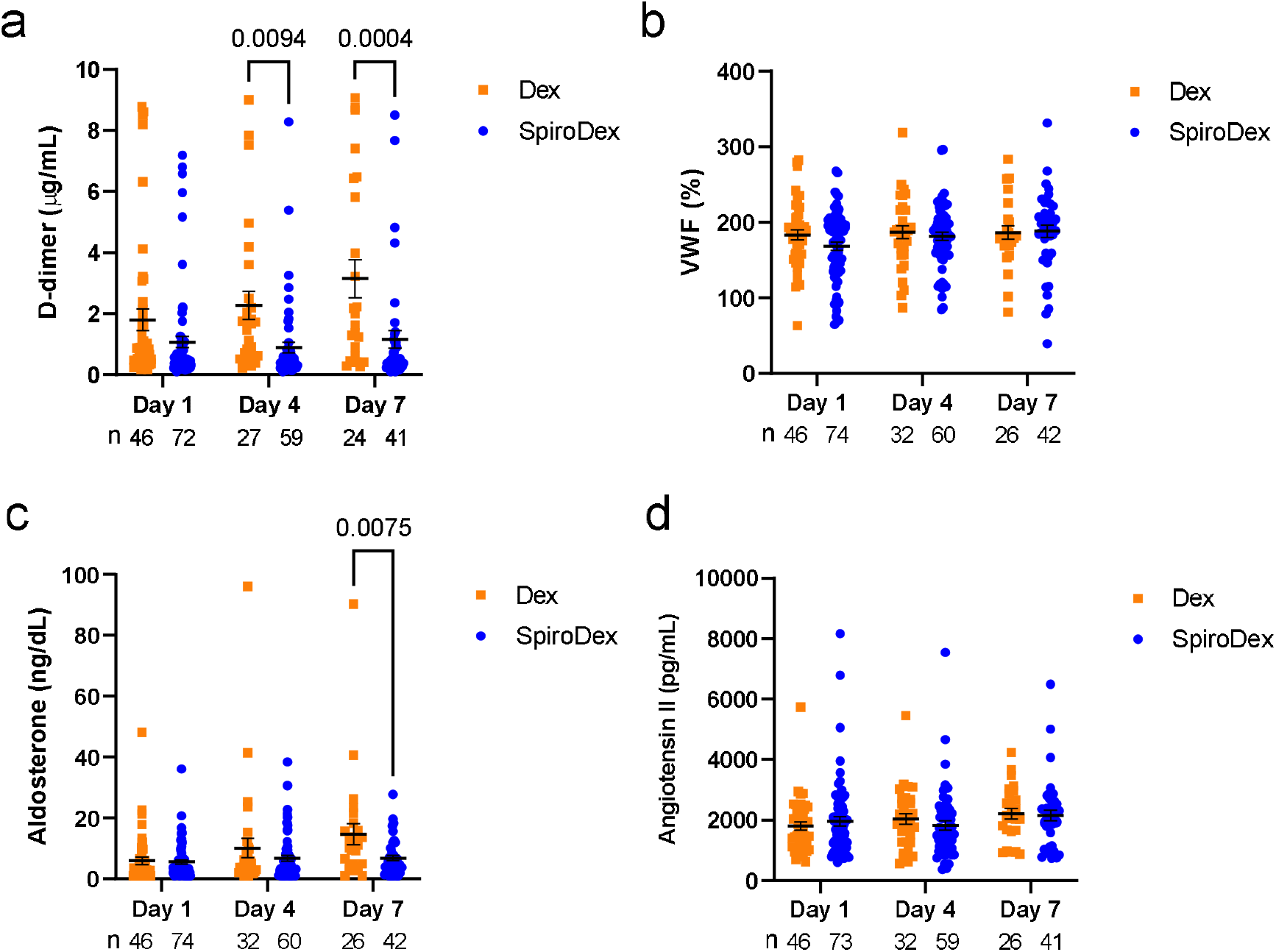
Primary Biomarker Outcomes. Day 1, 4 and 7 measurement of patient blood biomarkers **(a)** D-dimer, **(b)** VWF, **(c)** aldosterone and **(d)** angiotensin II. Mean and s.e.m shown. Analysed by two-way ANOVA, p < 0.05 indicated within the figure. For all, patient number (n) included in analysis is shown below the plot, Dex: squares, orange; SpiroDex: circles, blue.

### Secondary outcomes

There was no significant difference in the cough scores during the acute illness between the two groups (Figure 4a, b) however both day-time and night-time cough scores at day 28 were significantly lower in the SpiroDex group compared to the Dex group (Figure 4c, d) (D28 day-time: p = 0.037, D28 night-time p = 0.0136). There were no significant differences in plasma corticosteroid levels between groups (Figure 4e, f). Patients in the SpiroDex group reached oxygen freedom at a median of day 6 compared to a median of day 7 in the Dex group (Figure 4g), (p = 0.06).

**Figure 4.**
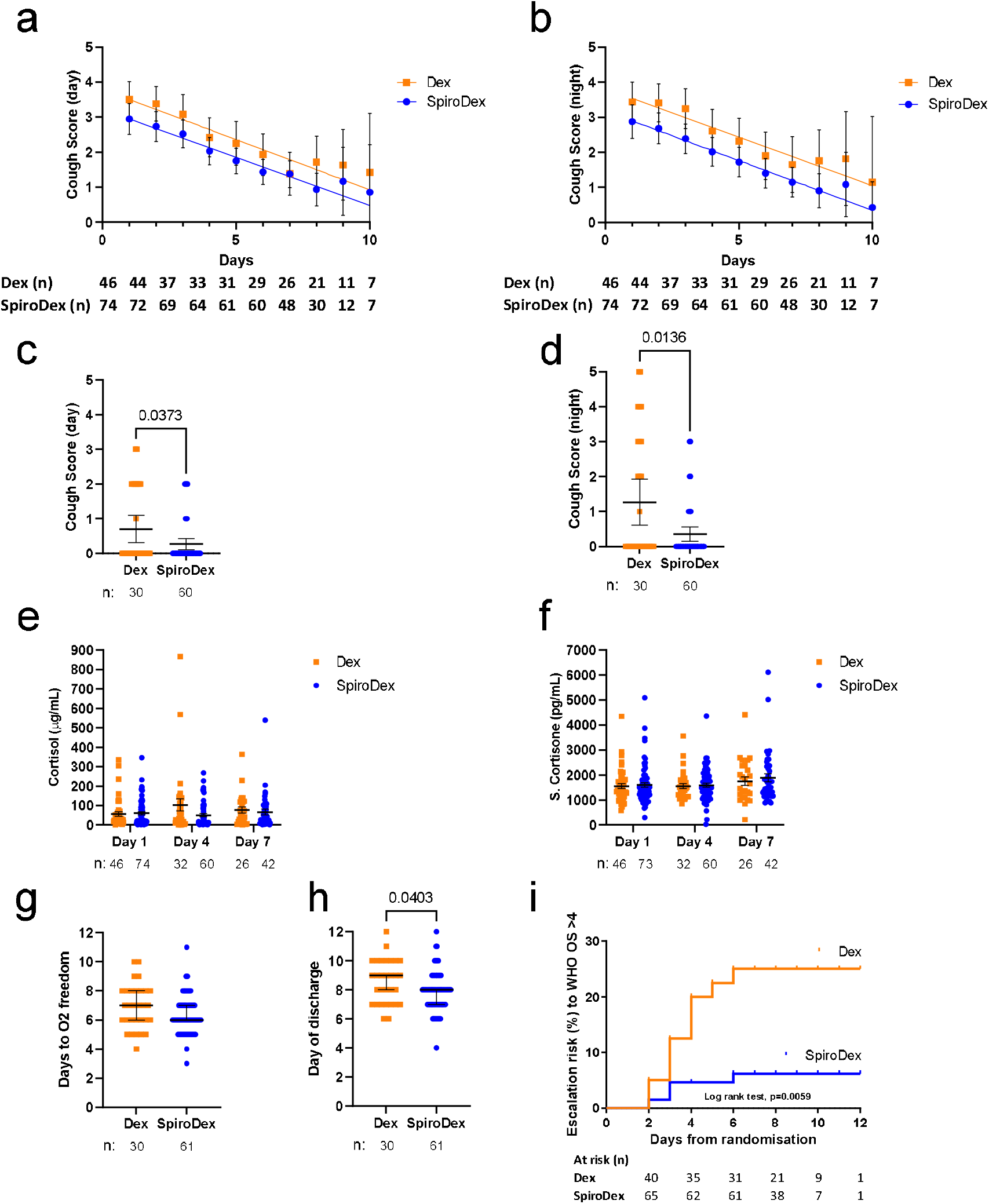
Secondary outcomes & post hoc analysis: Cough Score, oxygen use and plasma corticosteroid levels. **(a)** day-time and **(b)** night-time cough scores recorded on days 1-10. Range: 0 = no cough, 5 = distressing cough. Mean and 95% CI shown. Analysed by simple linear regression and two-way ANOVA. **(c)** Day 28 follow up day-time and **(d)** night-time cough score. Mean and 95% CI shown. Analysed by Mann-Whitney. Measurement of **(e)** plasma cortisol **(f)** and cortisone on days 1,4 and 7. Mean and s.e.m shown. Analysed by two-way ANOVA. **(g)** Days to supplemental oxygen freedom. Median and 95% CI shown. Analysed by Mann-Whitney. **(h)** Days to patient discharge. Median and 95% CI shown. Analysed by Mann-Whitney. **(i)** Kaplan Meier plot to show risk of patient escalating to WHO OS >4. Hazard ratio 0.21, 95% CI 0.070 to 0.64. For all, patient number (n) included in analysis is shown below the plot, p < 0.05 indicated within the figure. Dex: squares, orange; SpiroDex: circles, blue.

### Post hoc analysis

82.4% (n=61) of patients in the SpiroDex group were discharged from Lok Nayak Hospital (without reaching WHO OS scale >4), compared to 65.2% (n=30) of Dex patients. Of the patients who were discharged home, the SpiroDex patients were discharged earlier (median 8 days) than the Dex patients (median 9 days, p = 0.0403) (Figure 4h). Fewer patients in the SpiroDex arm compared to the Dex arm (5.4% vs 19.6%) were removed from the trial due to worsening COVID-19 (pre-specified criteria of escalating to WHO OS >4) (Figure 4i), demonstrated by a Kaplan Meier plot representing patients at risk of deteriorating to WHO OS >4.

### Adverse events

Low dose spironolactone as an-add on therapy to SoC was safe and well tolerated with no electrolyte differences between the groups (supp Figure S1). There were no significant adverse events (SAEs) reported. All patients received the full dose of Spironolactone throughout their inclusion in the trial and no patient required a reduced dose due to impaired renal function. Other clinical and safety biomarkers were similar across both groups (supp Figure S2). Aside from patients who were withdrawn due to worsening COVID-19 illness (Table S1) /escalation to critical care, AEs recorded were mild to moderate and self-limiting (Figure 1).

## Discussion

In this randomised placebo-controlled trial, hospitalised patients who received low dose spironolactone in addition to SoC had significantly lower D-dimer levels at days 4 and 7, and a lower aldosterone level at day 7. We did not observe a significant reduction in the time to recovery to WHO OS ≤ 3 but did observe a trend towards a shorter time to oxygen freedom and a significant reduction in day and night cough scores for SpiroDex patients at day 28.

Post hoc analysis demonstrated reduced clinical deterioration within the SpiroDex arm, and a reduced length of in-patient stay for those patients in the trial who were discharged from the Lok Nayak facility. In addition, treatment at this dose was safe with no recorded SAEs. This is the first report in a RCT setting of the potentially beneficial and safe addition of spironolactone to SoC in COVID-19.

We note several limitations to this trial. There was a higher proportion of male patients in the Dex than the SpiroDex group and it is known that males are at a higher risk of severe illness in COVID-19 (15). In addition, the SpiroDex group had statistically significant lower NEWS2 score and D-dimer levels and higher SpO2/FiO2 ratio at the time of recruitment (Table 1). The trial utilised manual randomisation with inherent limitations that comes with this method.

As the trial protocol did not allow for further data collection of patients after they deteriorated beyond WHO OS 4, the final outcomes for these individuals are unknown. In addition, a small number of patients had to be withdrawn from the trial due to their decision to be transferred to a different facility in Delhi. In total, 12.2% of patients within the SpiroDex group withdrew from the trial versus 15.2% from the Dex group.

A raised D-dimer is associated with increased mortality in COVID-19 disease, and it is a recognised prognostic indicator of adverse outcomes (16, 17). We observed a significantly reduced plasma D-dimer in the SpiroDex group at days 4 and 7 compared to the Dex group. This was also observed in a retrospective case series of MR antagonism (11), which reported a promising impact on mortality in patients with moderate to severe COVID-19, using intravenous canrenone, an MR antagonist. However, the dose of canrenone was dose equivalent to 120mg per day of spironolactone, as compared to 25mg used in our trial.

It is anticipated that SARS-CoV-2 will become endemic within the global population even with a successful vaccination program. Together with the potential of variants of the SARS-CoV-2 emerging, new affordable and widely available treatments are needed. The promising safety biomarker (D-dimer) and clinical outcomes demonstrated in this phase 2 RCT support the evaluation of spironolactone in COVID-19 in larger randomised phase 3 controlled trials.

## Data Availability

All data produced in the present study are available upon reasonable request to the authors.

## Acknowledgements

The trial in India was supported by the MAMC and the UK investigators were supported by the STOPCOVID project that has received a grant from LifeArc, an independent medical research charity. We would like to thank Dr Suri Labs Pvt Ltd for their assistance in biomarker evaluation.

## Author Contributions

BW & VM were chief investigators of the study and responsible for all trial related activities. BW, VM, SK, FH, NP, KS, VS contributed to the implementation of the study and all study related activities in Delhi. KF, AM, JA, AB, contributed to study support and resource allocation. TQ, LF analysed the blinded data. TQ, LF, BM, MS-H, CE and KD contributed to the interpretation of the data. All authors assisted with the writing of the paper and reviewed and approved the final manuscript.

## Supplemental data

**Table S1.** Further information on patients withdrawn from the trial due to worsening COVID19/ escalation to WHO Scale >4.

| Patient ID | Group | Co Morbid Conditions | Clinical status prior to exclusion from study |
| --- | --- | --- | --- |
| 073 | Dex | None. | Increased Oxygen requirement on day 2, SpO2/FiO2 86, NEWS 9, WHO scale 5. On Bilevel positive airway pressure non invasive ventilation (BiPAP) |
| 053 | Dex | Pulmonary Tuberculosis, Diabetes Mellitus | Increased Oxygen requirement on day 3, SpO2/FiO2 80, NEWS 9, WHO scale 5, Shifted to BiPAP |
| 097 | Dex | Hypertension, Diabetes Mellitus | Increased Oxygen requirement on day 3, SpO2/FiO2 78, NEWS 9, WHO scale 5, Shifted to BiPAP |
| 002 | Dex | Thrombocytopenia, Diabetes, Hyponatremia, external Hemorrhoids, Lung cancer with liver and brain metastasis | Reduced GCS with seizures: Intubated and put on mechanical ventilation |
| 016 | Dex |  | Increasing Oxygen demand on day 2, SpO2/FiO2 87, NEWS 9, WHO scale 5, Shifted to BiPAP |
| 023 | Dex | Diabetes, Hypertension CAD (2017): EF 35%, Diabetes Mellitus, ex-Smoker. Alcohol excess. | Deteriorated on Day 4 with increased oxygen demand, SpO2/FiO2 90, NEWS 9, WHO scale 5, Shifted to BiPAP |
| 060 | Dex | None. | Deteriorated on Day 2 with increased oxygen demand, SpO2/FiO2 87, NEWS 8, WHO scale 5, Shifted to BiPAP |
| 062 | Dex | Diabetes Mellitus, SARI-T1 Respiratory Failure, CAD | Deteriorated on Day 2 with increased oxygen demand, SpO2/FiO2 89, NEWS 7, WHO scale 5, Shifted to BiPAP |
| 088 | Dex | SARI, T1RF, Hypertension, Diabetes Mellitus | Increased Oxygen requirement on day 6, SpO2/FiO2 90, NEWS 8, WHO scale 5, Shifted to BiPAP |
| 008 | SpiroDex | Chronic Cough, Liver Abscess | Increased Oxygen requirement on day 2, SpO2/FiO2 88, NEWS 8, WHO scale 5<br>Shifted to BiPAP ventilation |
| 010 | SpiroDex | Hepatitis, Abdominal Pain, jaundice, Ventriculomegaly | Intubated and put on mechanical ventilation in view of poor GCS and increasing oxygen requirement |
| 117 | SpiroDex | Hypertension, Diabetes, Polio induced deformity | Increased Oxygen requirement on day 2, SpO2/FiO2 80, NEWS 9, WHO scale 5 put on BiPAP ventilation |
| 042 | SpiroDex | None. | Increased Oxygen requirement on day 2, SpO2/FiO2 78, NEWS 9, WHO scale 5, put on BiPAP |

**Figure S1.**
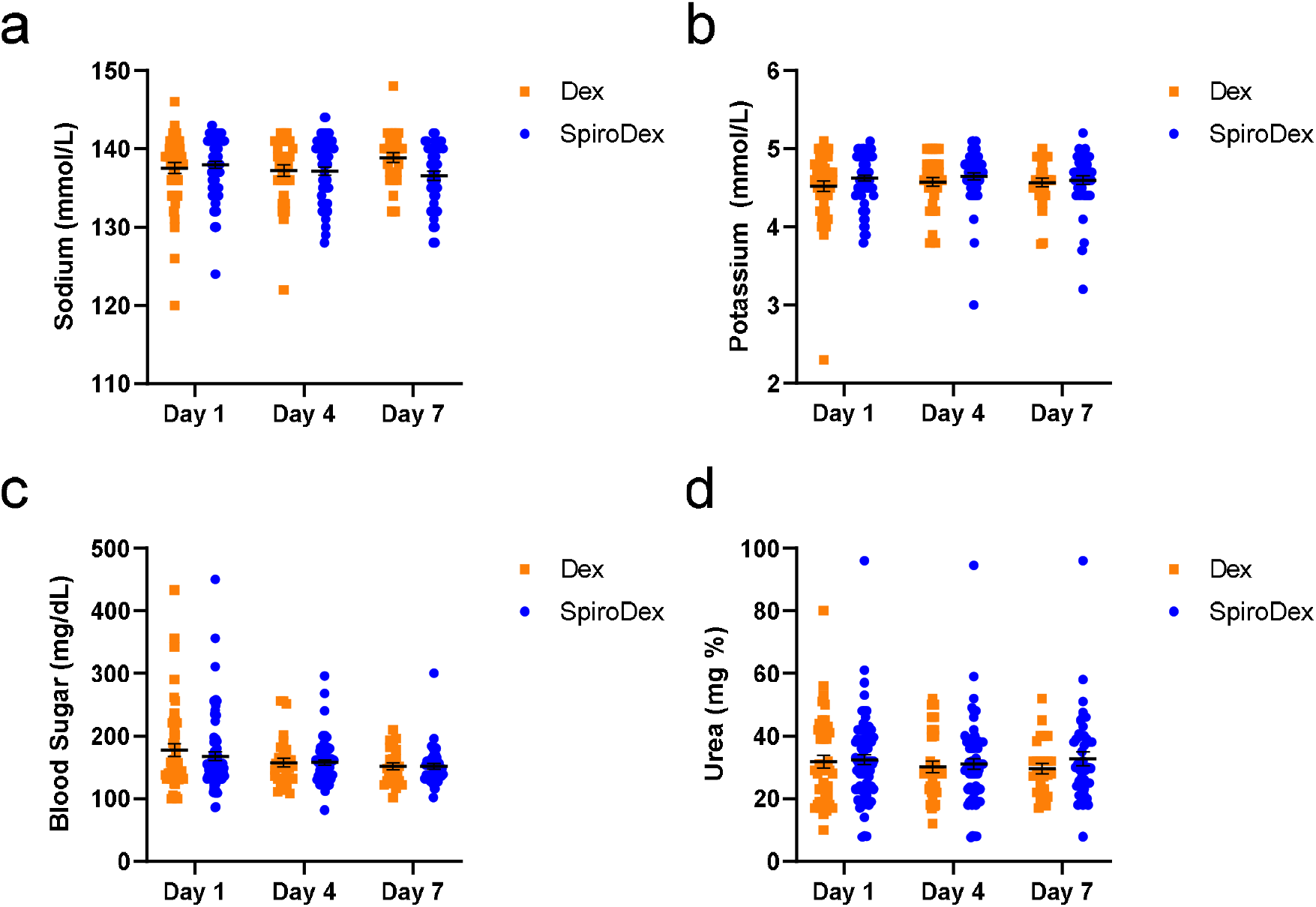
Additional clinical biomarkers. Day 1, 4 and 7 measurement of patient blood biomarkers **(a)** sodium, **(b)** potassium **(c)** blood sugar, and **(d)** Urea. Data shows mean and SEM. Dex: squares, orange; SpiroDex: circles, blue.

